# Factors associated with psychosocial distress among Rohingya refugees in Cox’s Bazar, Bangladesh: a representative household survey

**DOI:** 10.64898/2026.02.27.26347287

**Authors:** Harry J Wilson

## Abstract

The protracted Rohingya refugee crisis continues to deteriorate with approximately 1.2 million refugees currently sheltering in Cox’s Bazar, Bangladesh. This study aimed to estimate the prevalence and identify factors associated with psychosocial distress among Rohingya refugees. Data were sourced from the 2023 Joint Multi-Sectoral Needs Assessment - a representative cross-sectional household survey conducted across the 33 Rohingya refugee camps of Cox’s Bazar. Households were selected using stratified (by camp) random sampling. Psychosocial distress was assessed via proxy report by an adult household respondent and defined as the presence of at least one of eleven symptoms in the two weeks preceding the survey. Binary logistic regression was conducted to investigate household characteristics and individual factors associated with psychosocial distress status. The prevalence of psychosocial distress was 14.9% (95%CI: 14.1%–15.7%) among 16,455 Rohingya refugees sampled from 3,400 households. After adjustment, psychosocial distress was associated with individuals from aid-dependent households (aOR= 1.42 [95%CI: 1.21–1.67]), stress livelihood coping strategies (aOR= 3.03 [95%CI: 1.94–4.74]), crisis livelihood coping strategies (aOR= 4.40 [95%CI: 2.81–6.89]), emergency livelihood coping strategies (aOR= 4.15 [95%CI: 2.58–6.66]), individuals who required and received healthcare (aOR= 1.27 [95%CI: 1.12–1.43]), individuals who required and did not receive healthcare (aOR=1.49 [95%CI: 1.16–1.91]), individuals aged 18-34 years (aOR= 8.38 [95%CI: 6.99–10.04]), aged 35-59 years (aOR= 10.33 [95%CI: 8.44–12.65]), and aged 60+ years (aOR= 13.31 [95%CI: 10.25–17.30]). Psychosocial distress among Rohingya refugees was highly prevalent and associated with increasing age groups, aid dependency, negative livelihood coping strategies, and healthcare needs. The findings emphasise the need for comprehensive mental health and psychosocial support services in protracted humanitarian emergencies. Additional validation studies may be required to measure both the prevalence and severity of psychosocial distress to better inform humanitarian programming.

## Introduction

The mass displacement of the Rohingya - an ethnic minority from western Myanmar, has precipitated one of the most significant humanitarian crises in recent history [1,2]. Independent monitoring and verification activities conducted by the United Nations widely characterise the military persecution as acts of ethnic cleansing and bearing the hallmarks of genocide [1]. The mass influx of Rohingya fleeing targeted violence in late 2017 coalesced with previous waves of forced displacement, establishing one of the largest and most densely populated refugee settlements [3]. Among the 1,596,600 estimated refugees and asylum seekers from Myanmar, approximately three-quarters (1,178,000) are sheltering within the 33 refugee camps of Cox’s Bazar in neighbouring Bangladesh [4,5]. The Rohingya refugee crisis has transitioned from an acute emergency response to a protracted humanitarian setting, with refugees confined to deteriorating living conditions within the overcrowded camps, restrictions imposed on freedom of movement, limited educational access, scarce opportunities for employment, intensifying security concerns, and diminished prospects of dignified repatriation amid a resurgence in targeted violence across the border [3,6-8].

The psychological toll of targeted violence, forced displacement, statelessness and uncertainty experienced by refugees are well-established but poorly understood [9-19]. Current meta-analyses highlight the high prevalence of mental health disorders among refugees that substantially exceed the rates of depression, anxiety and post-traumatic stress disorder (PTSD) observed among the general population [12-15]. The ethnic violence and human rights violations endured by the Rohingya add an additional layer of complexity to the epidemiological profile of psychiatric illness among conflict-affected refugees [1,10,18]. Meta-analyses among Tutsi survivors of the 1994 Rwandan genocide highlight the elevated prevalence of post-traumatic stress disorder and the lengthy time-horizon from initial exposure to the emergence of psychiatric symptoms [10,17]. Additional evidence suggests that daily stressors endured by forcibly displaced communities - such as the everyday survival pressures that confront Rohingya refugee households within Cox’s Bazar, may exert equal or even greater influence on contemporary psychiatric symptomatology than prior exposure to traumatic events alone [11]. These programmatic implications are reinforced by concurrent demands for humanitarian relief in other sectors and prioritization of immediate physical health needs during acute emergency settings that often eclipse the provision of mental health and psychosocial support (MHPSS) services [2,8,20]. The lengthy time-horizon, prior exposure to historical trauma, contemporary survival pressures, and programmatic challenges in delivering specialized psychiatric services commonly result in the under-detection and inadequate management of highly prevalent psychiatric conditions among forcibly displaced refugee communities [9-19]. In conjunction, these factors underscore the need for proactive screening, early intervention, and prompt linkage to MHPSS services for Rohingya refugees to improve individual psychiatric outcomes, reduce psychosocial morbidity, and strengthen community resilience [9,19,21,22]. Despite the programmatic imperatives, representative evidence concerning the epidemiological profile of psychosocial distress among the Rohingya remains limited. This study aimed to estimate the prevalence and identify factors associated with psychosocial distress among Rohingya refugees living within the 33 camps of Cox’s Bazar, Bangladesh.

## Methods

### Study design and participants

This study utilised de-identified participant data from the 2023 Joint Multi-Sectoral Needs Assessment (J-MSNA), a representative cross-sectional household survey conducted across the 33 Rohingya refugee camps in Cox’s Bazar, Bangladesh [23]. The REACH Initiative coordinated the primary data collection in partnership with sectoral authorities and locally recruited enumerators [24]. The licensed dataset was obtained from UNHCR on the 27th of March 2025. All 191,271 Rohingya refugee households registered across the 33 camps of Cox’s Bazar were eligible for selection [24]. A comprehensive sampling frame of household addresses was constructed from administrative records maintained by camp authorities. Households were drawn using a stratified random sampling technique, with each camp (N=33) constituting a separate sampling stratum. The number of households sampled from each camp was determined by a single-proportion sample size formula that calculated the number of households required to ensure camp-specific estimates with a maximum 10% margin of error and using a 95% confidence level [24]. Finite population corrections were applied to compensate for the variable number of eligible households within each camp. The resultant sample sizes were inflated by 10% to mitigate anticipated non-response. Although the sampling strategy was principally designed to support camp-level household estimates to strategically inform humanitarian programming, the achieved sample size provides sufficient statistical precision for individual-level analyses using a rigorous sampling strategy generalisable at the response-level [20,25].

### Data Collection

Of the 3,465 households selected for sampling, 3,400 completed the survey between 27 August and 17 September 2023, with individual data pertaining to 18,172 household members. Data collection was conducted via face-to-face household interviews using a standardised structured questionnaire designed to capture multi-sectoral humanitarian indicators. Gender-balanced enumerator teams administered interviews to an adult household member (aged 18 years or older) matched by self-identified gender, who provided proxy responses on behalf of all individuals within the household. Enumerators recorded responses electronically via the KoBo Collect application and uploaded participant data to a secure UNHCR server. Raw participant data were cleaned for quality assurance purposes in accordance with the REACH Initiative minimum standards. A final de-identified dataset was procured for analysis from cleaned participant records. Verbal informed consent was obtained from each household respondent and documented within the final dataset. Prior to participation, respondents were informed of the survey purpose, assured of response confidentiality, and advised of their right to refuse any question or discontinue the interview without consequence. Enumerators received training in participant safeguarding principles and were equipped with referral protocols for any urgent protection or health concerns identified during data collection. Records with missing data for any variable were minimal (<1%) and were excluded from the analysis. While efforts were made to link case numbers recorded at the household level with individuals within the household for statistical analysis, no efforts were made to identify anonymous participants and all results remain de-identified.

### Variables

The primary outcome was psychosocial distress status assigned to individual household members according to proxy responses from an adult participant interviewed by enumerators. Psychosocial distress was defined as the presence of at least one of eleven symptoms experienced within the two weeks preceding the survey. These symptoms included: (i) nightmares, (ii) lasting sadness, (iii) extreme fatigue without doing work, (iv) being often tearful, (v) hopeless for the future, (vi) avoiding people, places or activities due to feelings of distress, (vii) anxiety or extreme worry for the future, (viii) extreme anger and out of control, (ix) uninterested in things that they used to like, (x) unable to carry out essential activities, and (xi) changes in appetite or sleep pattern compared to usual. The conceptualisation of psychosocial distress as an umbrella term encompassing a broad spectrum of emotional, cognitive, behavioural, and somatic manifestations follows the Inter-Agency Standing Committee (IASC) guidelines on MHPSS in emergencies [9,16]. Unlike discrete psychiatric conditions and their specific diagnostic criteria, psychosocial distress describes a cluster of related symptoms linked to depression, anxiety, and post-traumatic stress disorder (PTSD) that are often experienced by refugees in response to displacement, trauma, and ongoing adversity in protracted crisis settings [9,15,16,21,26]. The specific symptoms used to ascertain the presence of psychosocial distress during J-MSNA surveys are drawn from the WHO-UNHCR Assessment Schedule of Severe Symptoms in Humanitarian Settings (WASSS), Kessler Psychological Distress Scale, and Patient Health Questionnaire items [22,27,28].

The majority of psychosocial distress cases (80.3%) reported by adult respondents uniquely identified specific members within the household by their age-group (3-17, 18-59, 60+) and gender (male, female) characteristics. The remaining 19.7% of cases that were not uniquely identifiable were randomly assigned among 1,088 eligible members matched for the same age-group (3-17, 18-59, 60+) and gender (male, female) characteristics of those reported to be experiencing psychosocial distress within the household by the proxy respondent. To strengthen robustness, this process of random selection was repeated during sensitivity analysis as described in the statistical analysis section to inform variance and validate the main results. The random selection of ambiguous cases supported by sensitivity analysis was preferred over listwise deletion at the household level which favoured smaller households with fewer members, or listwise deletion at the individual level which favoured older household members that were more likely to be directly assigned case status. Additionally, the assignment of case status at the individual level allowed for individual factors (such as health needs by care status) to be adjusted for during multivariate analysis rather than restricting the investigation to only household variables and individual gender-specific age-group categories (3-17, 18-59, 60+) used to record case status among household members.

Individual variables and household characteristics evaluated for association with psychosocial distress status were identified from previous J-MSNA research, Refugee Influx Emergency Vulnerability Assessments (REVA), and funding impact analyses conducted by the World Food Programme [29-32]. Disability status was classified as a binary variable according to participants reporting either “a lot of difficulty” or “cannot do it at all” in any of the six functional domains as per the Washington Group Short Set (WG-SS) guidelines [33]. Household livelihood coping strategies were categorised as none, stress, crisis, or emergency as per World Food Programme guidelines [34]. Stress livelihood coping strategies included spending savings to meet essential needs, buying food on credit or borrowing food, borrowing money to meet essential needs, selling household assets/goods, selling or exchanging food assistance, and selling non-food items that were provided as assistance to meet other essential needs [34]. Crisis strategies included selling productive assets or means of transport (sewing machine, wheelbarrow, bicycle etc.), reducing essential non-food expenditure (education, health, clothing etc.), or withdrawing children from school to meet essential needs [34]. Emergency strategies included children (below 18 years) working to contribute to household income, adults (18 years and older) accepting high risk illegal income activities, begging or scavenging to meet essential needs, household migration, marriage of any children (below 18 years) as they had become a financial burden, and marriage of a female child under 18 years to intermix with the local population hoping to secure a more stable lifestyle [34].

### Statistical analysis

All results were computed using Stata version 19.5 survey functions that accounted for the stratified sampling design at the camp-level, adjusted for the clustering of individuals within the same household, and controlled for differential probability of household selection between the camps (strata) using survey weights computed as the inverse probability of selection [35].

Descriptive statistics were computed to analyse the characteristics of sampled households and household members. Survey-weighted frequencies and survey-weighted percentages were reported for categorical variables. Survey-weighted median and survey-weighted interquartile range (IQR) were reported for asymmetrical continuous variables. Survey-weighted mean and survey-weighted standard deviation (SD) were reported for symmetrical continuous variables. Survey-weighted percentages and 95% confidence intervals (95%CIs) were reported for prevalence estimates.

Binary logistic regression analysis was performed to investigate household and individual factors associated with the odds of psychosocial distress. Unadjusted odds ratios (OR) and adjusted odds ratios (aOR) were reported for bivariate and multivariate estimates respectively. A threshold value of p ≤ 0.05 was used to conclude statistical significance and 95%CIs were reported for precision using design-adjusted standard errors calculated via the Taylor series linearization method.

A sensitivity analysis using random selection (488 cases among 1,088 eligible individuals) with repeated sampling (2,000 repeats) was conducted to evaluate how the random assignment of non-identifiable cases (n=488) among eligible household members (N=1,088) influenced the logistic regression estimates. Unadjusted and adjusted estimates were repeated across the 2,000 unique samples. Histograms were plotted to confirm normality of point estimates and standard errors. The mean point estimate and mean standard error from the sensitivity analysis were contrasted with their equivalents from the main results to identify any problematic distortion induced by the random selection process. The mean point estimate and mean standard error were used to construct 95%CIs, and cross-validated with the OR (95%CI) and aOR (95%CI) estimates from the main analysis. The main results pertain to a single cohort (cohort 1 of 2,000 used during sensitivity analysis) and are presented for simplicity as justified by confirmatory results observed during sensitivity analysis. The odds attributed to individual health needs by care status were evaluated for potential problematic criterion overlap by contrasting estimates after excluding individuals that sought care for MHPSS services. Pairwise correlation and variance inflation factor (VIF) analyses were conducted to evaluate potential problematic multicollinearity. Receiver operator characteristics (ROC) and area under the curve (AUC) analyses were conducted to assess the multivariate model’s ability to accurately discriminate psychosocial distress status. AUC estimates were repeated as part of the sensitivity analysis and model performance was evaluated using the mean AUC with 95%CI reported for precision.

## Results

### Participant characteristics

Of the 18,172 household members surveyed, 1,717 were aged under 3 years and excluded from subsequent analysis. The median age was 18 years (IQR: 10 **–** 30; Table 1) among the remaining 16,455 individuals sampled from 3,400 households. The proportion of children aged 3-17 (47.4%) and adults aged 18-59 (46.9%) was approximately equal, with the remaining 5.7% contributed by elderly household members aged 60 or older. Slightly more females (50.9%) were sampled compared to males (49.1%), and the mean household size was 5 members (SD= ±2). Over a third of households contained at least one member in psychosocial distress (38%), with most reporting either one member (14.8%) or two members (17.4%) in psychosocial distress. Households with three or more members in psychosocial distress were relatively infrequent (5.8%).

**Table 1.** Characteristics of surveyed household members aged 3+ (N= 16,455) and sampled households (N= 3,400).

|  | Summary* |
| --- | --- |
| **Age, median** | 18 [IQR: 10 – 30] |
| Children (3-17) | 7,791 (47.4%) |
| 3-11 | 5,133 (31.2%) |
| 12-17 | 2,658 (16.2%) |
| Adults (18-59) | 7,719 (46.9%) |
| 18-29 | 4,147 (25.2%) |
| 30-44 | 2,293 (13.9%) |
| 45-59 | 1,279 (7.8%) |
| Elderly (60+) | 945 (5.7%) |
| **Gender** |  |
| Male | 8,082 (49.1%) |
| Female | 8,373 (50.9%) |
| **Household Size, mean (N= 3,400)** | 5 (±2) |
| 1-3 | 1,032 (30.4%) |
| 4-6 | 1,653 (48.6%) |
| 7+ | 715 (21.0%) |
| **Prevalence of Psychosocial Distress** | 14.9% (95% CI: 14.1%–15.7%) |
| **Members in Psychosocial Distress per Household (N= 3,400)** |  |
| None | 2,108 (62.0%) |
| One | 503 (14.8%) |
| Two | 592 (17.4%) |
| Three or More | 197 (5.8%) |

\* Summary statistics are weighted frequencies and weighted percentages for categorical variables, weighted median (IQR) for asymmetrically distributed continuous variables, weighted mean (SD) for symmetrical continuous variables, or weighted population prevalence (%) and 95% CIs for outcome variables.

### Factors associated with psychosocial distress

Table 2 shows the logistic regression results investigating factors associated with the odds of psychosocial distress. In bivariate analysis, psychosocial distress was associated with head of household characteristics (gender, age, marital and disability status), household income sources, utilisation of livelihood coping strategies, individual disability status, healthcare needs, individual age and gender. After adjustment in multivariate analysis, psychosocial distress was associated with individuals from aid-dependent households (aOR= 1.42 [95%CI: 1.21–1.67]), stress livelihood coping strategies (aOR= 3.03 [95%CI: 1.94–4.74]), crisis livelihood coping strategies (aOR= 4.40 [95%CI: 2.81–6.89]), emergency livelihood coping strategies (aOR= 4.15 [95%CI: 2.58–6.66]), those who required and received healthcare (aOR= 1.27 [95%CI: 1.12–1.43]), those who required and did not receive healthcare (aOR=1.49 [95%CI: 1.16–1.91]), individuals aged 18-34 years (aOR= 8.38 [95%CI: 6.99–10.04]), aged 35-59 years (aOR= 10.33 [95%CI: 8.44–12.65]), and aged 60+ years (aOR= 13.31 [95%CI: 10.25–17.30]).

**Table 2.** Factors associated with the odds of psychosocial distress.

|  | Summary | Odds Associated with Psychosocial Distress |  |  |  |
| --- | --- | --- | --- | --- | --- |
|  | (N= 16,455) | Unadjusted |  | Adjusted |  |
|  | n (%) | OR (95%CI) | p-value | aOR (95%CI) | p-value |
| **Head of Household Gender** |  |  |  |  |  |
| Male | 13,534 (82.2%) | 1.00 | – | 1.00 | – |
| Female | 2,921 (17.8%) | **1.30 (1.12–1.51)** | **<0.001\*** | 1.05 (0.84–1.31) | 0.666 |
| **Head of Household Age Group** |  |  |  |  |  |
| 18-24 | 874 (5.3%) | 1.00 | – | 1.00 | – |
| 25-34 | 4,327 (26.3%) | 0.87 (0.66–1.14) | 0.317 | 1.20 (0.89–1.61) | 0.239 |
| 35-44 | 4,174 (25.4%) | 0.88 (0.66–1.16) | 0.355 | 1.28 (0.94–1.74) | 0.123 |
| 45-59 | 4,229 (25.7%) | 0.78 (0.59–1.04) | 0.088 | 0.87 (0.63–1.19) | 0.383 |
| 60+ | 2,851 (17.3%) | 1.19 (0.89–1.59) | 0.240 | 0.95 (0.68–1.31) | 0.736 |
| **Head of Household Marital Status** |  |  |  |  |  |
| Married | 14,753 (89.7%) | 1.00 | – | 1.00 | – |
| Divorced/Separated/Single/Widow | 1,702 (10.3%) | **1.39 (1.16–1.67)** | **<0.001\*** | 1.21 (0.93–1.57) | 0.148 |
| **Head of Household Disability Status** |  |  |  |  |  |
| No Disability | 13,472 (81.9%) | 1.00 | – | 1.00 | – |
| Living with a Disability | 2,983 (18.1%) | **1.39 (1.18–1.64)** | **<0.001\*** | 1.14 (0.92–1.42) | 0.231 |
| **Income Source** |  |  |  |  |  |
| Income Source | 12,307 (74.8%) | 1.00 | – | 1.00 | – |
| No Income (Aid-Dependent) | 4,148 (25.2%) | **1.48 (1.29–1.70)** | **<0.001\*** | **1.42 (1.21–1.67)** | **<0.001\*** |
| **Household Livelihood Coping Strategies** |  |  |  |  |  |
| None | 977 (5.9%) | 1.00 | – | 1.00 | – |
| Stress | 7,431 (45.2%) | **2.89 (1.90–4.41)** | **<0.001\*** | **3.03 (1.94–4.74)** | **<0.001\*** |
| Crisis | 5,772 (35.1%) | **3.79 (2.48–5.78)** | **<0.001\*** | **4.40 (2.81–6.89)** | **<0.001\*** |
| Emergency | 2,275 (13.8%) | **3.55 (2.28–5.54)** | **<0.001\*** | **4.15 (2.58–6.66)** | **<0.001\*** |
| **Individual Disability Status** |  |  |  |  |  |
| No Disability | 15,734 (95.6%) | 1.00 | – | 1.00 | – |
| Living with a Disability | 721 (4.4%) | **2.43 (2.01–2.95)** | **<0.001\*** | 1.14 (0.90–1.44) | 0.286 |
| **Individual Health Need(s) and Care Status** |  |  |  |  |  |
| No Health Need | 7,008 (42.6%) | 1.00 | – | 1.00 | – |
| Needed and Received Healthcare | 8,442 (51.3%) | **1.57 (1.40–1.76)** | **<0.001\*** | **1.27 (1.12–1.43)** | **<0.001\*** |
| Needed and Did Not Receive Healthcare | 1,005 (6.1%) | **2.09 (1.68–2.61)** | **<0.001\*** | **1.49 (1.16–1.91)** | **0.002\*** |
| **Individual Age-Group** |  |  |  |  |  |
| 3-17 | 7,791 (47.4%) | 1.00 | – | 1.00 | – |
| 18-34 | 5,102 (31.0%) | **7.71 (6.41–9.27)** | **<0.001\*** | **8.38 (6.99–10.04)** | **<0.001\*** |
| 35-59 | 2,617 (15.9%) | **9.87 (8.20–11.90)** | **<0.001\*** | **10.33 (8.44–12.65)** | **<0.001\*** |
| 60+ | 945 (5.7%) | **13.23 (10.48–16.71)** | **<0.001\*** | **13.31 (10.25–17.30)** | **<0.001\*** |
| **Individual Gender** |  |  |  |  |  |
| Male | 8,082 (49.1%) | 1.00 | – | 1.00 | – |
| Female | 8,373 (50.9%) | **1.08 (1.01–1.15)** | **0.017\*** | 0.98 (0.92–1.05) | 0.570 |

\* Statistically significant at $p \leq 0.05$ .

Sensitivity analysis demonstrated strong alignment between the main results and the mean results from random selection of cases that could not be perfectly assigned with 2,000 repeats. All point estimates (odds ratios) and standard errors from unadjusted and adjusted logistic regression were within ±2%. All variables were cross-validated with the corresponding estimates from sensitivity analysis using the mean point estimate and mean standard error to construct 95%CIs to ensure robustness of results presented in Table 2. No problematic criterion overlap was observed after removing participants that sought care for MHPSS services with point estimates from unadjusted and adjusted analysis within ±4%. The AUC indicated good discriminatory ability to distinguish between individual psychosocial distress status (AUC= 0.770 [95%CI: 0.769–0.772]; Figure 1). No evidence of problematic multicollinearity was observed, all pairwise correlations between independent variables were below 0.7 and the mean VIF was 2.4 with no extreme outliers (VIF_max_ <10).

**Fig 1.**
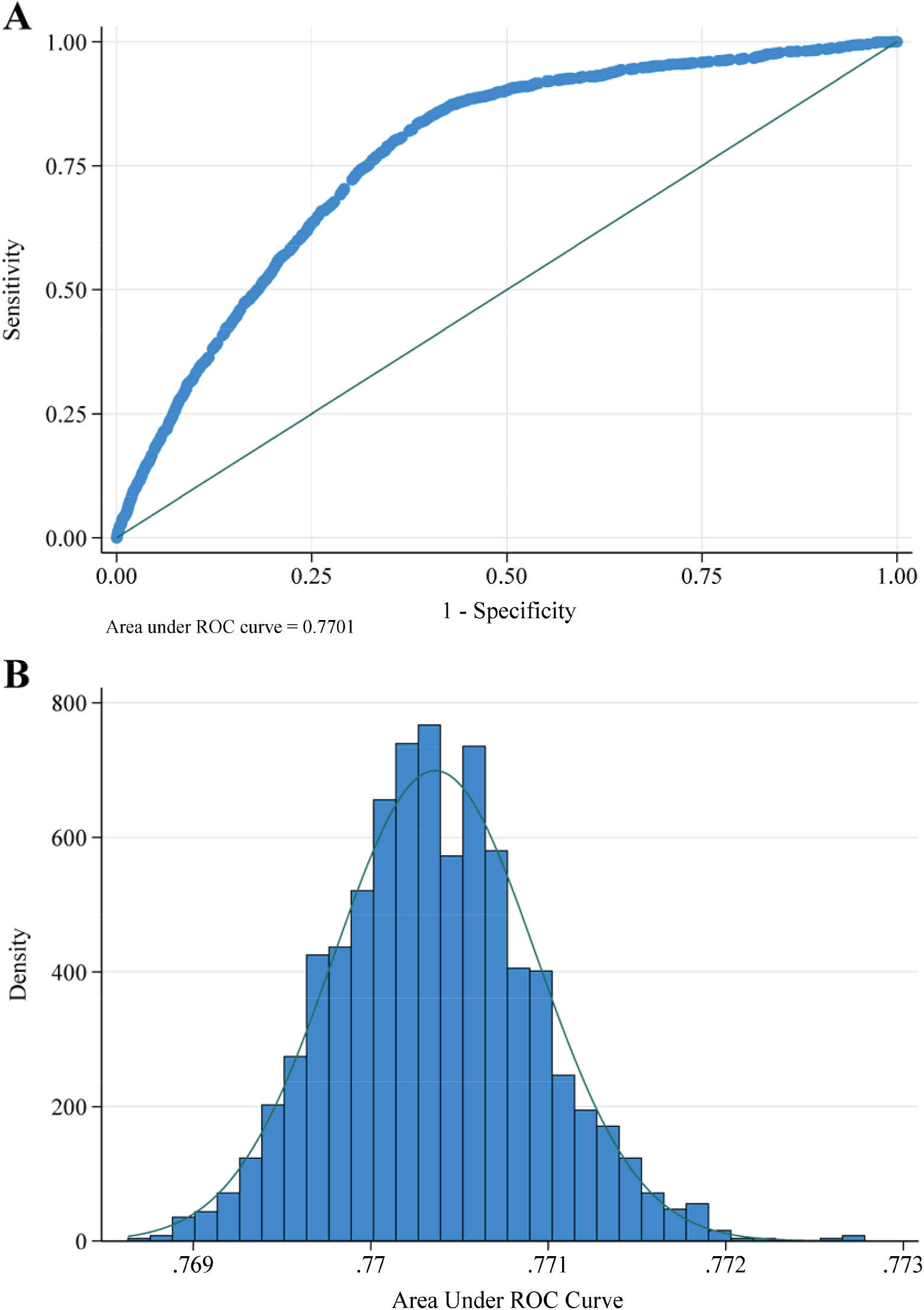
**A)**. Receiver operator characteristics (ROC) and area under the curve analysis (AUC) evaluating the discriminatory ability of the logistic regression model to distinguish between individual psychosocial distress status. **B)**. Histogram illustrating the distribution of area under the curve (AUC) values during sensitivity analysis (random selection with 2,000 repeats).

## Discussion

The sociodemographic profile of sampled participants strongly aligns with the population characteristics of Rohingya refugees in Cox’s Bazar as informed by UNHCR’s database [4,20]. The rigorous sampling method, face-to-face data collection, and alignment of demographic characteristics strongly support the representativeness of participants and hence generalisability of the results. The results highlight that approximately 15% of Rohingya refugees (age 3+) experienced symptoms of psychosocial distress in the preceding fortnight and over a third of households contained at least one member in psychosocial distress. These prevalence estimates are consistent with updated results from the 2024 Inter-Sector Needs Assessment (ISNA) which found that 32% of households contained at least one member experiencing emotional distress or trauma, and 39% of households contained a member who received psychological or mental health support [7].

After adjustment for potential confounding in multivariate analysis, increasing age-groups, aid dependency, utilisation of negative livelihood coping strategies, and healthcare needs by care status were significantly associated with the odds of psychosocial distress status. Specific unadjusted results such as female headed households, single headed households, and disability status, warrant further consideration rather than dismissed as spurious. Despite the study design limitations that preclude inferences of how these variables may be arranged within causal pathways, these findings evoke a cautious acknowledgment of how survival pressures disproportionately influence disadvantaged cohorts to adopt negative livelihood coping strategies or abstain from healthcare services they require but cannot necessarily afford – particularly among aid-dependent households. In this scenario, these particular variables that demonstrated significance in unadjusted analysis but found to be non-significant in multivariate analysis may reflect overadjustment bias from mediated effects rather than adjustment for confounding effects acting outside of a potential causal pathway.

The discontinuity of ordinal effects associated with crisis and emergency livelihood coping strategies requires methodological consideration and careful interpretation. The ordinal scale of livelihood coping strategies measure severity at the household level, whereas the broad operationalisation of psychosocial distress in humanitarian settings captures the occurrence of any eleven common symptoms. As such, the strength of association from regressing an ordinal independent variable upon a broad binary outcome are sensitive to the number of members in psychosocial distress within the household rather than the severity of distress they experience. In consequence, deriving inferences from the comparative strength of association between crisis and emergency livelihood coping strategies are problematic as they may reflect a measurement ceiling effect where all members within the household are experiencing psychosocial distress [36]. This proposed ceiling effect would explain the ordinal odds associated with increasing severity of livelihood coping strategies across the none, stress and crisis categories, as well as the comparable estimates observed between crisis and emergency categories due to the measurement ceiling. The minor attenuation observed may also be attributable to members who may have been in psychosocial distress but have left the household due to the emergency livelihood coping strategies involving underage marriage [34]. Reconceptualising the scale of psychosocial distress to measure severity among individuals rather than the number of members experiencing symptoms of psychosocial distress within the household may be required to prevent measurement ceiling and other scale attenuation issues. In the context of humanitarian needs analysis, measuring the severity of psychosocial distress through a surrogate of symptom-specific frequency rather than binary occurrence should be considered if operationally appropriate and strategically beneficial to inform the delivery of MHPSS services.

The results should also be interpreted in light of additional limitations. The random assignment of psychosocial distress status for a minority of cases that could not be perfectly assigned naturally induces a degree of misclassification bias. However, the results observed during sensitivity analysis demonstrated limited variance of estimates regardless of individual case status among eligible members within the household. The ascertainment of psychosocial distress status through proxy respondents at the household level rather than individual self-reported data may be less sensitive at detecting symptoms of psychosocial distress not readily apparent to the proxy respondent. This source of measurement bias is further exacerbated by the two-week time horizon that introduces recall bias, as proxy respondents were required to recount psychosocial distress symptoms among individual household members over the past two weeks. The cross-sectional design prevents causal inferences and the results may be prone to unmeasured confounding – especially at the individual level where limited data were captured from proxy respondents.

## Conclusion

Despite the notable study limitations, the findings offer valuable insights concerning the high prevalence and factors associated with psychosocial distress among Rohingya refugees in Cox’s Bazar. The analysis revealed that increasing age-groups, aid dependency, negative livelihood coping strategies, and healthcare needs were independently associated with individual psychosocial distress status. The pronounced age gradient and elevated odds associated with elderly refugees underscore the cumulative psychological toll of historical events, forced displacement, targeted violence, and trauma exacerbated by the diminished prospects of dignified repatriation. The findings have implications for targeted programming and emphasise the need for comprehensive mental health and psychosocial support services in protracted humanitarian emergencies. Additional validation studies may be required to certify methods used to measure both the prevalence and severity of psychosocial distress. Further longitudinal studies should be considered to elucidate the arrangement of variables that may act within causal pathways to influence individual psychosocial distress status.

## Data Availability

The de-identified participant-level dataset underlying the study results may be available upon reasonable request to UNHCR at https://microdata.unhcr.org/index.php/catalog/1128 subject to approval based on their data sharing policies, ethical requirements, and data use agreements.

## Acknowledgements

The author wishes to formally acknowledge those whose collective efforts have made this research article possible. Specifically, the Inter-Sector Coordination Group (ISCG), UNHCR, IOM, ECHO, the REACH Initiative, enumerators, other study personnel, and household respondents who graciously donated their time.

## Supporting information

**S1 Document**. STROBE checklist for cross-sectional studies.

